# Inflammatory signature and restriction of adaptive immunity are associated with unfavorable outcomes on immune checkpoint blockade in patients with advanced head and neck squamous cell carcinoma

**DOI:** 10.1101/2024.11.29.24317276

**Authors:** Lisa Paschold, Christoph Schultheiss, Paul Schmidt-Barbo, Konrad Klinghammer, Dennis Hahn, Mareike Tometten, Philippe Schafhausen, Markus Blaurock, Anna Brandt, Ingunn Westgaard, Simone Kowoll, Alexander Stein, Axel Hinke, Mascha Binder

## Abstract

**Background:** In most patients with relapsed or metastatic head and neck squamous cell carcinoma (rmHNSCC), immunotherapy with PD-1 targeting antibodies does not yield durable responses. PD-L1 tissue expression - the most commonly assessed marker for checkpoint inhibiting antibodies - is an insufficient predictor of treatment outcome.

**Methods:** We evaluated various blood and tissue-based biomarkers in the context of immune checkpoint blockade-based treatment to find suitable response biomarkers in a clinical trial cohort of patients with rmHNSCC.

**Results:** The PD-L1 expression level in tumor or tumor microenvironment was not associated with treatment benefit. In contrast, inflammation-related markers such as IL-6, high peripheral neutrophils and high levels of cell-free DNA, as well as markers related to adaptive immune dysfunction such as altered T cell dynamics and secretion of immune checkpoint molecules, were associated with poor clinical outcomes. Patients lacking these high-risk markers performed remarkably well on inhibition of immune checkpoints with pembrolizumab.

**Conclusions:** Biomarker-guided patient selection for pembrolizumab monotherapy or novel combinatorial approaches - potentially including anti-inflammatory agents – for patients with immune-impaired, inflammatory profiles may be the next step in personalizing immunotherapy for these hard-to-treat patients.

## Background

Head and neck squamous cell carcinomas (HNSCC), primarily originating from the squamous epithelium of the oral cavity, pharynx and larynx, rank as the sixth most prevalent cancer globally, with nearly 900,000 new cases and approximately 450,00 deaths per year (1-4). HNSCC predominantly affect males and is causally linked to tobacco and alcohol consumption as well as infection with human papillomavirus (HPV) strains 16 and 18 (1, 2, 5-7). Early-stage locoregional disease can often be cured, but more than half of these cases relapse and around 15-30% develop metastatic disease (rmHNSCC) (1, 8, 9). Survival time for patients rmHNSCC has doubled over the past decade, primarily due to advances in systemic treatment (1, 4, 10-12). Despite this progress, the median overall survival (OS) for these patients remains limited to 12-14 months (12, 13).

Preclinical data indicate that HNSCC is highly immunosuppressive, marked by abnormal proinflammatory cytokine secretion and impaired immune effector cell function (14, 15). Therefore, one of the major advances in systemic treatment of rmHNSCC was the introduction of immunotherapies. Landmark phase III trials have led to the approval of two anti-programmed cell death-1 (PD-1) antibodies - pembrolizumab and nivolumab - for use in this indication (16, 17). More recently, pembrolizumab also received approval as a first-line treatment with or without concomittant chemotherapy (18). Despite these advancements, only a subset of patients with rmHNSCC benefit from immunotherapy, underscoring the urgent need to identify novel biomarkers to optimize treatment strategies. PD-L1 is currently the only predictive biomarker for response to immune checkpoint inhibitors in rmHNSCC in routine clinical practice, but there is still an important subset of patients with PD-L1 positive disease that does not derive benefit from these drugs (19, 20).

In this study, we conducted comprehensive biomarker analyses using biomaterial from patients enrolled in the FOCUS trial, all of received pembrolizumab. Our objective was to identify simple, clinically applicable markers to predict the outcome of checkpoint inhibitor treatment. Our findings demonstrate that patients with a high-inflammation and impaired T cell profile experience suboptimal outcomes with immune checkpoint inhibition alone, indicating the need for additional or alternative therapeutic approaches.

## Results

### Patient characteristics, study treatment and survival outcomes in the cohort

The trial enrolled 75 evaluable patients from August 2021 to July 2023. 25 were enrolled into the calibration arm receiving pembrolizumab monotherapy and 50 into an experimental arm receiving pembrolizumab in combination with the hTERT vaccine UV1. Detailed patient characteristics are described in (21). The study population was representative for patients with rmHNSCC in that it also included 18% of patients with an ECOG performance score of 2.

The trial did not meet its primary endpoint. UV1 treatment did not result in higher-than-expected rate of progression-free survival (PFS) at six months. Therefore, we combined both pembrolizumab-containing study arms for the biomarker analysis shown here. To avoid any potential biases from the experimental UV1 vaccination, all analyzes were additionally plotted by treatment arm as shown in the supplementary data section. PFS and overall survival (OS) is shown in Figure 1A and B. With a median follow-up of 11.3 months, the median PFS and OS were very comparable to the pembrolizumab arm of the KEYNOTE-048 trial that had enrolled only patients up to an ECOG performance score of 1 (18). The median PFS of 3.4 months compared to 2.3 months for patients with PD-L1 CPS 1 or more and 3.4 months for patients with PD-L1 CPS 20 or more in the KEYNOTE-048 trial (18). The median OS of 13.1 months compared to 13.0 months in the KEYNOTE-048 trial (18).

**Figure 1:**
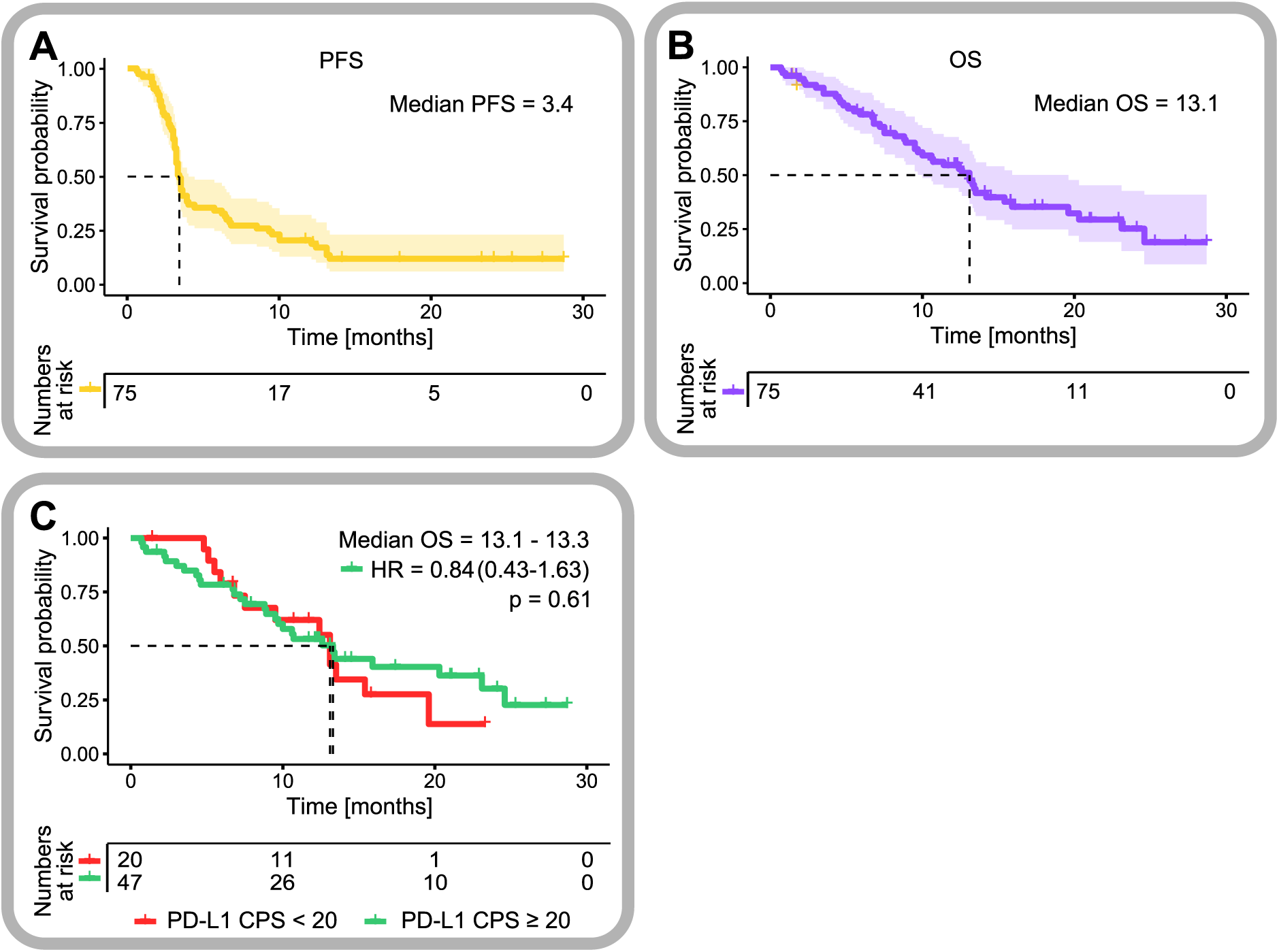
Efficacy of pembrolizumab (+/- UV1) treatment in patients with R/M HNSCC and as a function of PD-L1 CPS in the FOCUS trial. Kaplan-Meier estimates of progression-free (A) and overall survival (B) and according to PD-L1 CPS subgroup (cut-off 20) (C). HR = hazard ratio. Statistic test = log rank test.

### Efficacy as a function of PD-L1 CPS

PD-L1 expression is an established prognostic biomarker in HNSCC (22, 23). Interestingly, in the PD-L1 positive population studied in the FOCUS trial, high or low PD-L1 combined positivity scores (PD-L1 CPS below or above 20) did not appear to segregate patient subsets with higher or lower clinical benefit (Figure 1C and Supplementary Figure 1A and B).

### T cell receptor repertoire profiling

Previous studies from our group had shown that T cell repertoire metrics may be a strong predictive biomarker for immune checkpoint blockade in distinct disease settings (24-27). We assessed T cell immune repertoires by next-generation sequencing of the T cell receptor beta (TRB) locus from patient blood at BL and prior to the second pembrolizumab dose in both arms. Patient immune repertoires showed differences in global T cell immune metrics to healthy age-matched control individuals (Figure 2A). While BL T cell metrics did not appear to determine outcomes on pembrolizumab (data not shown), the dynamics of T cell repertoire clonality correlated with overall survival. Patients with increasing T cell repertoire restriction (increase of T cell repertoire clonality >20% above BL) showed unfavorable outcomes on pembrolizumab compared to patients with stable repertoires (Figure 2B and Supplementary Figure 2A and B).

**Figure 2:**
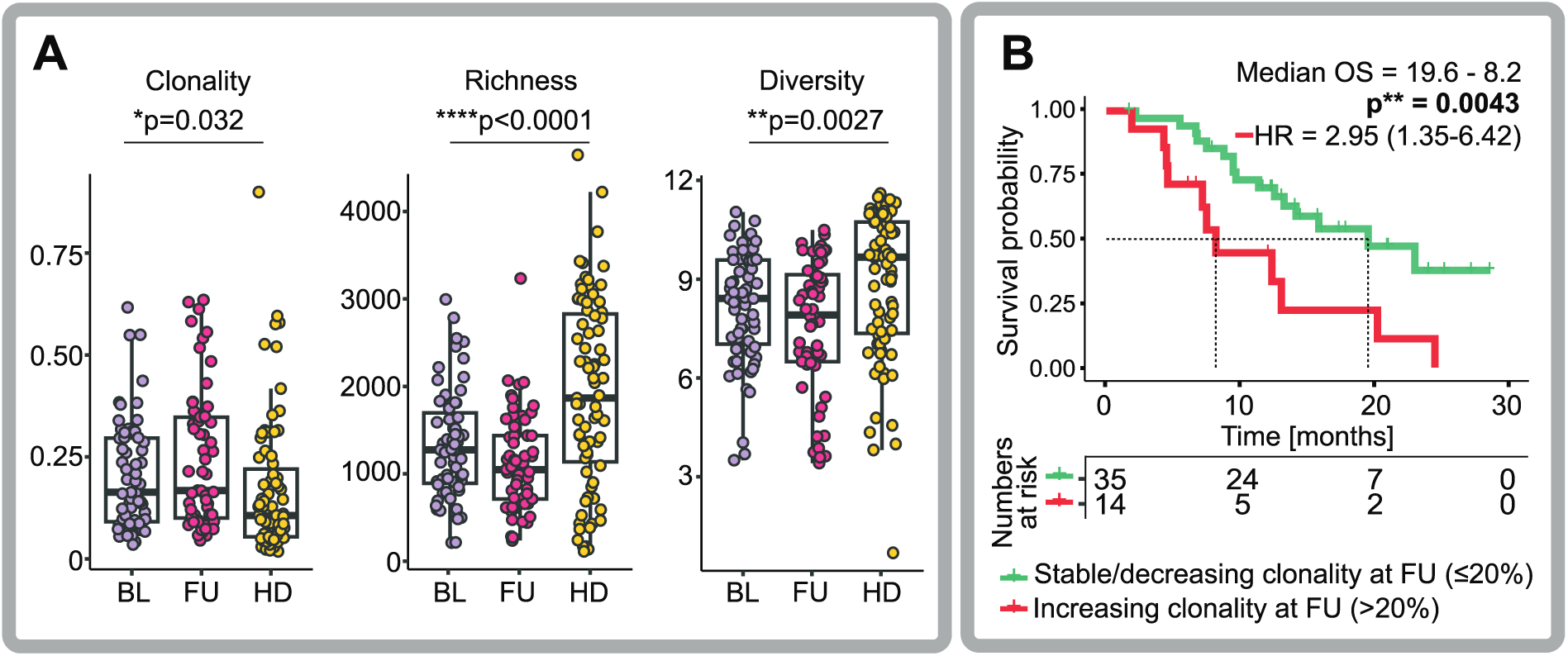
T cell receptor repertoire metrics in HNSCC patients treated with pembrolizumab on the FOCUS trial. **(A)** Blood T cell repertoire clonality, richness and diversity in healthy individuals (HD) and the HNSCC patient population of the FOCUS trial and healthy control individuals are shown at baseline assessment (BL) and before the second treatment cycle (follow-up; FU). **(B)** Overall survival (OS) outcomes of patients with increasing T cell restriction (>20% increase in T cell receptor clonality over the first pembrolizumab treatment cycle). HR = hazard ratio. Statistic test = log rank test.

### Blood soluble factor analysis

HNSCC is marked by abnormal proinflammatory cytokine secretion (28). As a proxy of the degree of inflammation, we assessed cytokine patterns in the blood of our patients. Moreover, we were interested in the levels of circulating checkpoint molecules some of which had been found to counteract immune checkpoint inhibition in other disease settings (29, 30). Blood testing was performed at baseline (BL) and before the second treatment cycle (follow-up, FU). As shown in Figure 3A and 3B, most of the analyzed inflammatory factors displayed high plasma levels at both sampling time points as compared to healthy individuals. Notable exceptions were IL- 1β, IL-17A, IFN-γ and GM-CSF (Figure 3A). While mean levels of TNF, IL-6 and IL-23 trended towards lower levels at FU, IL-10, IFN-β and IP-10 showed the opposite pattern (Figure 3A). IFN-λ1 and IFN-α2 were only elevated at FU but not at BL time points (Figure 3A).

**Figure 3:**
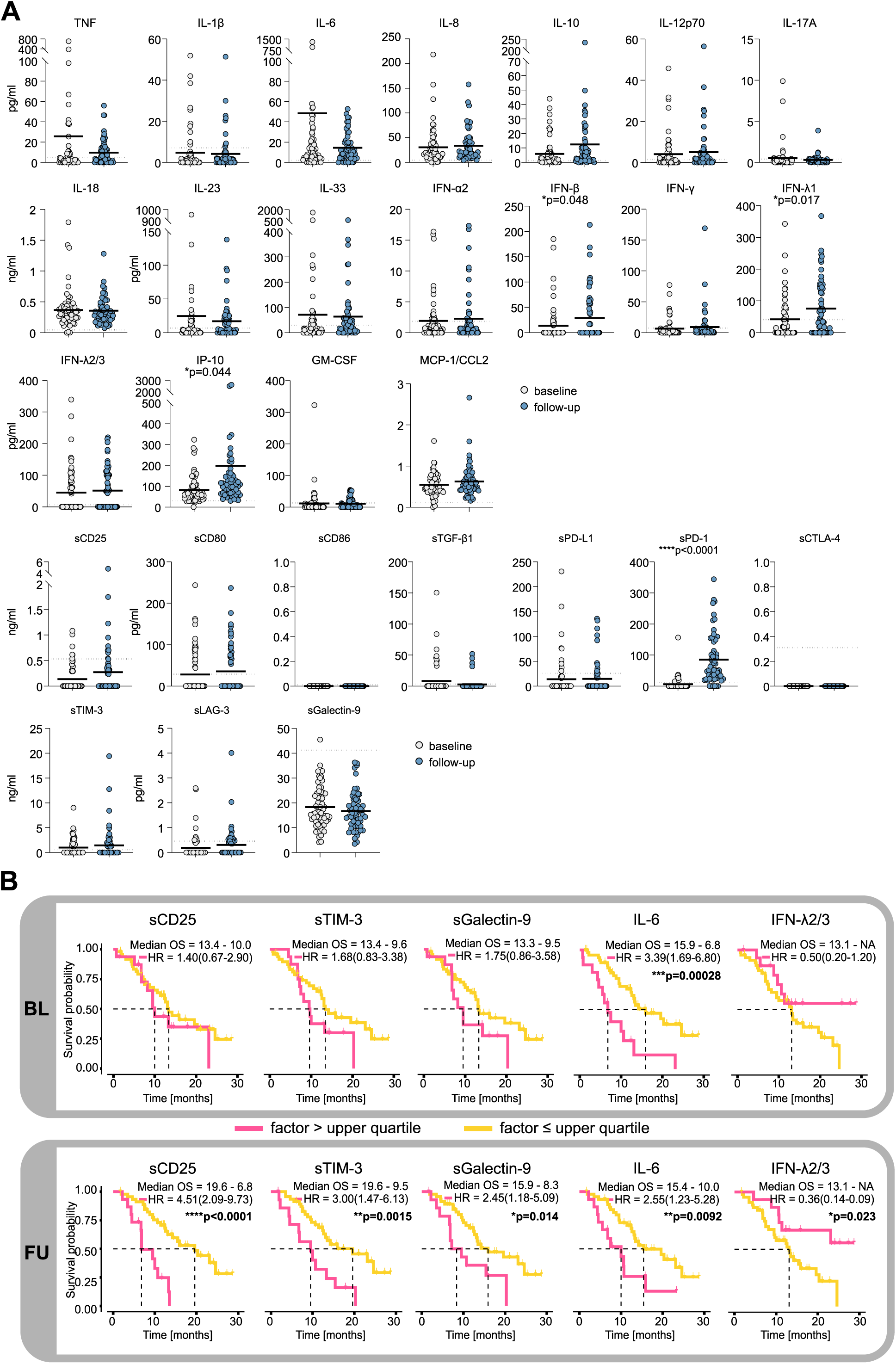
Soluble factor and chemokine profiling baseline and upon pembrolizumab treatment. **(A)** Circulating immune checkpoint molecules and cytokines in patients from the FOCUS trial at baseline assessment (BL) and before the second cycle of pembrolizumab (follow-up; FU). Dashed lines indicate means from 20 randomly chosen healthy donors. **(B)** Overall survival (OS) outcomes on pembrolizumab in patients with elevated levels of circulating immune checkpoint molecules and cytokines (upper quartile) as compared to the rest of the cohort at BL and on treatment. HR = hazard ratio. Statistic test = log rank test.

In contrast, plasma levels of most soluble immune checkpoints were actually diminished compared to the levels measured in 20 healthy blood donors (sCD25, sPD-L1, sCTLA-4, sLAG-3 and sGalectin-9; Figure 3B). Exceptions were sTIM-3 which was slightly elevated at both time points and sPD-1 which was highly elevated after only one dose of the PD1-directed checkpoint inhibitor at FU (Figure 3A). Next, we assessed the clinical courses in patients with high levels of selected soluble factors. Individuals with levels in the upper quartile were considered high level. When testing these patients against the rest, we observed that patients with high cytokine plasma levels had a lower survival probability on pembrolizumab as compared to patients with lower levels (Figure 3B). This was especially true for patients with elevated levels of sCD25, sTIM-3, IL-6 and sGalectin-9 and more pronounced after the first dose of the immune checkpoint inhibitor (FU) than at BL assessment (Figure 3B; Supplementary Figure 3A and B). Interestingly, this was reversed in patients with high IFN-λ2/3 levels (Figure 3B).

### Cell-free DNA levels and neutrophil to lymphocyte ratios (NLR)

In the inflammatory context of cancer, high levels of DNA seem to be released into the blood resulting in elevated cell-free DNA (cfDNA) levels in cancer patients as compared to healthy individuals. The majority (roughly 75%) of this DNA has been found to derive from neutrophils rather than from the tumor itself or its immediate environment (31). Given the inflammatory signature of HNSCC in general and the unfavorable outcomes on immune checkpoint inhibition observed with this signature in our FOCUS trial, we wished to analyze cfDNA levels along with the NLR as potentially predictive biomarkers. We reasoned that lower cfDNA levels and lower NLR may define a patient subpopulation that might benefit from pembrolizumab monotherapy without the need for additional e.g. chemotherapy.

cfDNA levels of normal individuals have been shown to have cfDNA levels around 4.3 ng/ml, while patients with stage I-III cancer have levels around 12. 6 ng/ml (31). In line with this, our patients had elevated cfDNA levels with a median of 11.9 ng/ml (Figure 4A). Also, NLR were elevated in our cohort (median of 6.18) as compared to healthy individuals that typically have a ratio of 1-3 (Figure 4B). Patients with cfDNA in the lower quartile of our cohort (below 7.9 ng/ml) and/or NLR in the lower quartile (below 4.1) had favorable clinical courses on pembrolizumab (Figure 4C and Supplementary Figure 4A and B). Yet, these two features – cfDNA levels and NLR - did not show any obvious correlation (Table 1).

**Figure 4:**
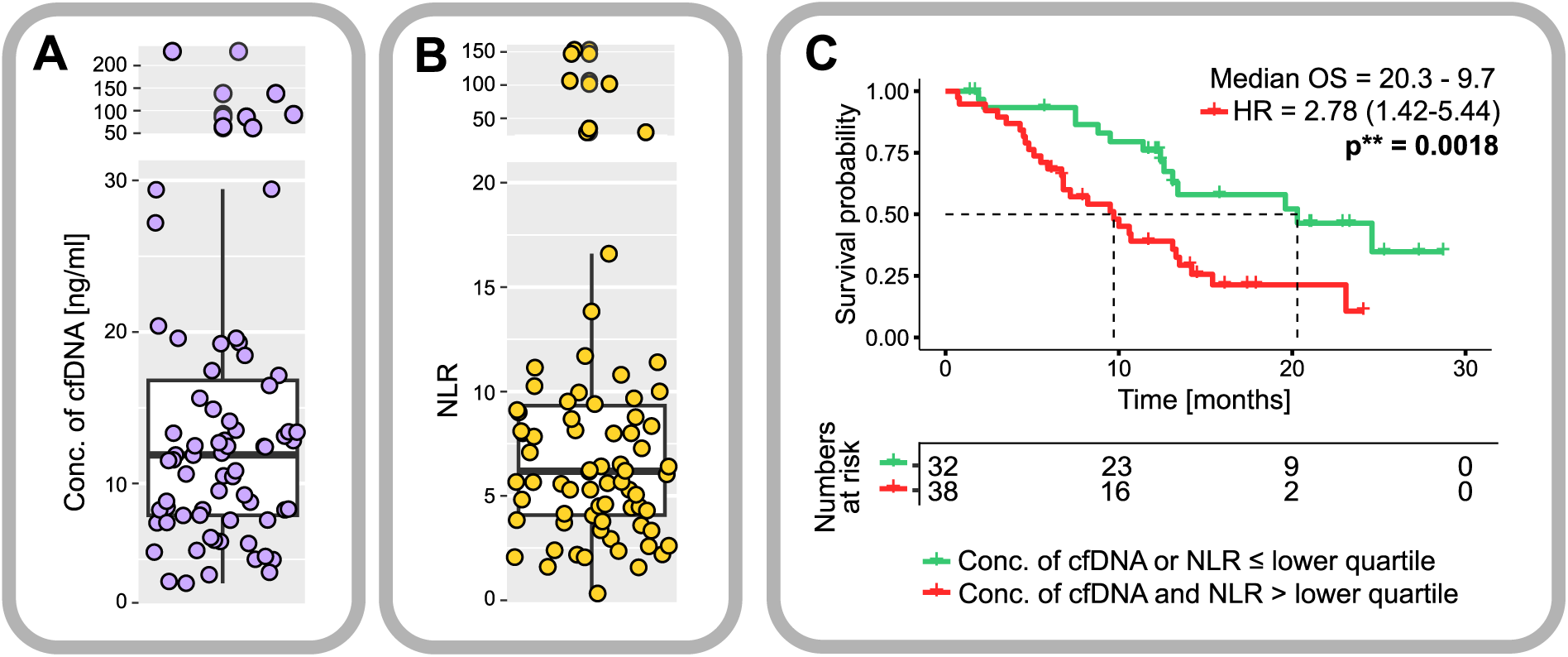
Cell-free DNA (cfDNA) and neutrophil-to-lymphocyte ratios (NLR) in patients with HNSCC treated with pembrolizumab on the FOCUS trial. **(A)** cfDNA levels and **(B)** NLR in HNSCC patients from the FOCUS trial at baseline assessment. **(C)** Overall survival (OS) in patients with low cfDNA and/or NLR (lower quartile). HR = hazard ratio. Statistic test = log rank test.

**Table 1:** Patient subsets from the FOCUS trial displaying high or low cfDNA and/or NLR at baseline assessment.

|  | cfDNA high | cfDNA low |  |
| --- | --- | --- | --- |
| NLR high | 38 | 15 | 53 |
| NLR low | 15 | 2 | 17 |
|  | 53 | 17 | 70 |

### Immunotypes and biomarker importance tested in the FOCUS trial

We performed a broader unsupervised cluster analysis to understand potential immunotypes beyond individual markers. All biomarkers that had shown correlation with OS in the individual analyzes were included. We identified essentially three clusters (Figure 5A): One group of patients was mainly characterized by secretion of the soluble immune molecules sTim3 and sCD25 at FU (red), another group showed high levels of IFN-λ2/3 at FU (green), and a third group showed neither of these features (violet). Overall survival was short in the sTim3/sCD25 group, long in the IFN-λ2/3 group and intermediate in the group without these features (Figure 5B). A comprehensive analysis including all of these biomarkers suggested a positive correlation of the sTim3/sCD25 cluster with high sGalectin-9 and – to a lesser extent – with sPD-L1 levels (Figure 5C). There was some correlation of sPD-L1 levels with IL-6 levels measured at FU. All other biomarkers tested did not show significant correlation with this or other clusters.

**Figure 5:**
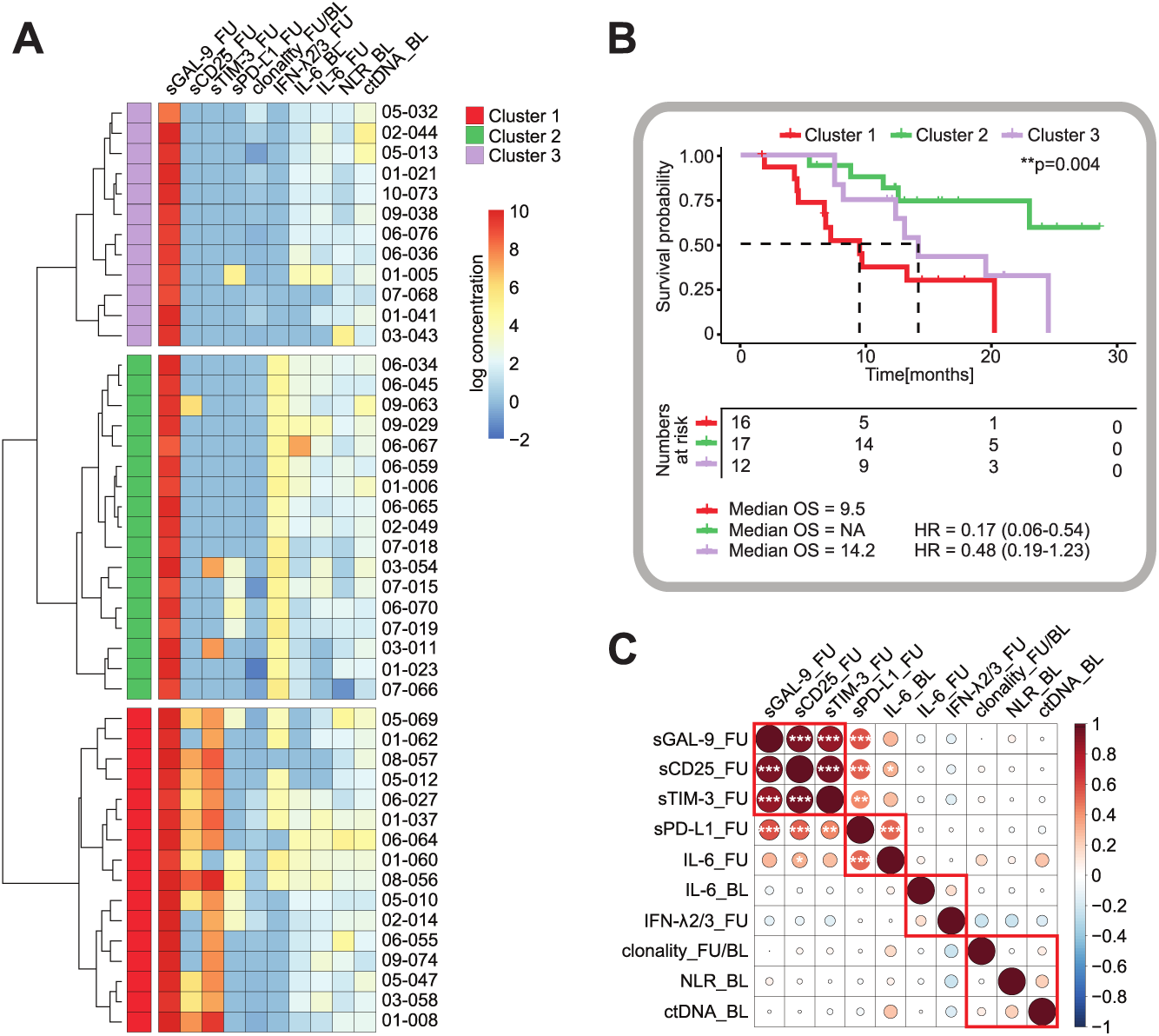
Unsupervised cluster analysis to determine immunotypes of patients with HNSCC on pembrolizumab treatment. **(A)** Unsupervised hierarchical clustering of log10 transformed cytokine concentrations in plasma samples of indicated patients on the FOCUS trial. **(B)** Survival analysis for the three patient clusters as established in (A). OS = overall survival, HR = hazard ratio. Statistic test = log rank test. **(C)** Correlation of parameters studied in (A) in a correlation matrix. Asterisks indicate significant correlations.

## Discussion

Immune checkpoint inhibitors have significantly expanded the treatment options for rmHNSCC. However, the ideal treatment schedules and combination partners are still uncertain. In fit patients with relapsed or refractory PD-L1-positive HNSCC, pembrolizumab as a monotherapy or in combination with chemotherapy has demonstrated comparable effectiveness in broad, unselected cohorts (13, 17). It is likely that specific patient subgroups benefit more from the addition of chemotherapy, while others may achieve sufficient responses with pembrolizumab alone. Furthermore, certain patients may require completely different therapeutic approaches altogether. This question is critical, as head and neck cancer patients often present with comorbidities, and the addition of chemotherapy can significantly increase treatment-related toxicities.

In our study, we treated a cohort of patients with up to and including ECOG 2 performance status using pembrolizumab. In the study-arm B, patients received an hTERT vaccine in addition to the pembrolizumab backbone, but this combination failed to meet the primary endpoint of PFS and was therefore deemed ineffective (21). Nonetheless, we identified several intriguing biomarkers in this cohort that correlate with clinical outcomes. These markers were associated with inflammatory processes (e.g., elevated IL-6 levels, high cfDNA concentrations, increased neutrophil-to-lymphocyte ratios) or impaired T cell immunity (e.g., high T cell repertoire clonality, secretion of immune checkpoint molecules). Patients exhibiting a pronounced inflammatory profile or restricted T cell repertoire with high immune checkpoint molecule secretion experienced poor outcomes on pembrolizumab. However, due to the study design, it remains unclear whether these factors are predictive or merely prognostic.

Moreover, the role of inflammation in HNSCC as a resistance mechanism to immunotherapy, or as a direct driver of tumor progression, remains unresolved (14, 15). This observation raises the important question of whether combining immunotherapy with chemotherapy or other agents, such as anti-inflammatory drugs, could enhance treatment efficacy and improve patient outcomes. One potential approach could involve the combination of immune checkpoint inhibitors with JAK inhibitors, which target signaling pathways downstream of interferon and type 1 cytokine receptors on immune cells. Recent studies have shown success with this concept in non-small cell lung cancer and Hodgkin’s lymphoma (32, 33). In these models, JAK inhibition with ruxolitinib or itacitinib reversed CD8+ T cell exhaustion driven by myeloid-derived suppressor cell (MDSC)-mediated IFN-JAK/STAT signaling. While MDSCs are also present in HNSCC tumors, their infiltration is not yet linked to disease etiology, though infiltration rates do correlate with clinical stage (34). Notably, elevated NLRs have been associated with higher MDSC frequencies and worse prognosis across several cancer types (35-38).

Furthermore, JAK inhibition could offer the added benefit of direct anti-tumor activity, given the reliance of many HNSCC tumors on JAK-STAT signaling pathways. This dual-action approach - targeting both immune modulation and tumor signaling - holds promise for improving outcomes in this challenging patient population.

Taken together, these findings suggest that rmHNSCC subsets with dismal outcomes on immune checkpoint inhibition as monotherapy may be identifyable by blood-based diagnostics. Refining immunotherapeutic strategies for these patients, possibly through combination with chemotherapy or other anti-inflammatory agents, may overcome resistance mechanisms and ultimately improve outcomes in rmHNSCC.

## Methods

### Biomaterial from the FOCUS clinical trial

Patients donated 20 mL peripheral blood (collected in STRECK cell-free DNA BCT tubes) at baseline assessment and prior to the second pembrolizumab dose for translational research.

### Survival analysis

Kaplan-Meier estimates were calculated based on cox models with p-values derived from log rank test and hazard ratios derived from cox regressions. Missing values were removed. All comparative tests should be considered exploratory. All analyses and plots were generated with R version 4.3.3, RStudio version 2024.12.1 and packages survminer (39) and survival (40).

### Next-generation T cell receptor repertoire sequencing

Leukocytes were pelleted from STRECK tubes and genomic DNA was isolated using the GenElute mammalian genomic DNA miniprep kit (Sigma-Aldrich, Taufkirchen, Germany) according to the manufacturer’s instructions. To analyze blood immune cells throughout treatment, amplification of the T cell receptor beta chain (TRB) repertoire from circulating cells was performed as described elsewhere (25, 41-46). Sequencing and de-multiplexing was performed on the Illumina MiSeq platform with 2 x 301 cycles at an average coverage of 80,000 reads per sample. Analysis of the TRB locus were performed using MiXCR V3.0.12 (47). All analyses and data plotting were carried out using R version 4.3.3 (48) and the package tcR (49). (13). Student’s t-test was used to compare two groups and ANOVA was used to evaluate multiple groups. All comparative tests should be considered exploratory. The datasets generated in this study has been deposited in the European Nucleotide Archive (ENA, ID: PRJEB80902).

### cfDNA quantification

Isolation of cfDNA from blood plasma using was performed using the QIAamp Circulating Nucleic Acid Kit (Qiagen, Hilden, Germany) and quantified using Qubit dsDNA high-sensitivity assay (Thermo Fisher Scientific, Waltham, USA)

### Soluble factor analysis

Plasma cytokines and other soluble factors were quantified using the LEGENDplex Human Immune Checkpoint Panel (10-plex) and the Human Inflammation Panel (13-plex) (BioLegend) according to the manufacturer’s instructions. Read out of the LEGENDplex assays was performed on a CytoFLEX flow cytometer (Beckman Coulter Life Science). Correlation of plasma levels were calculated using the R package corrplot with the R version 4.3.and RStudio 2023.06.1.

## DECLARATIONS

### Contributors

MBi and AS designed the study/research program. LP and CS performed experiments. PSB and LP performed bioinformatical analyses. AB, KK, DH, MT, PS, MBl, SK, AS recruited patients and/or provided technical and material support. AH performed statistical analyses. MBi, LP, CS, PSB, AB, AS and AH analyzed and interpreted the data. MBi, LP and CS wrote the initial draft of the manuscript. All authors edited and approved the manuscript.

## Acknowledgements

We thank all patients and families as well as all participating study centers.

## Data availability statement

Data are available in the European Nucleotide Archive (ENA, ID: PRJEB80902) or upon request.

## Ethics statements

This study is based on participants of the FOCUS trial (NCT05075122), which was conducted at 10 centers in Germany under approval of the local ethics committees and in compliance with the Declaration of Helsinki. All participants provided written informed consent.

## Abbreviations

BL: baseline
cfDNA: cell-free DNA
FU: follow-up
MDSC: myeloid-derived suppressor cell
NLR: neutrophil to lymphocyte ratios
OS: overall survival
PD-1: programmed cell death-1
PD-L1: programmed cell death ligand-1
PFS: progression-free survival
rmHNSCC: relapsed or metastatic head and neck squamous cell carcinoma
TRB: T cell receptor beta

