## Supplemental Information for "Inflammatory signature and restriction of adaptive immunity are associated with unfavorable outcomes on immune checkpoint blockade in patients with advanced head and neck squamous cell carcinoma"

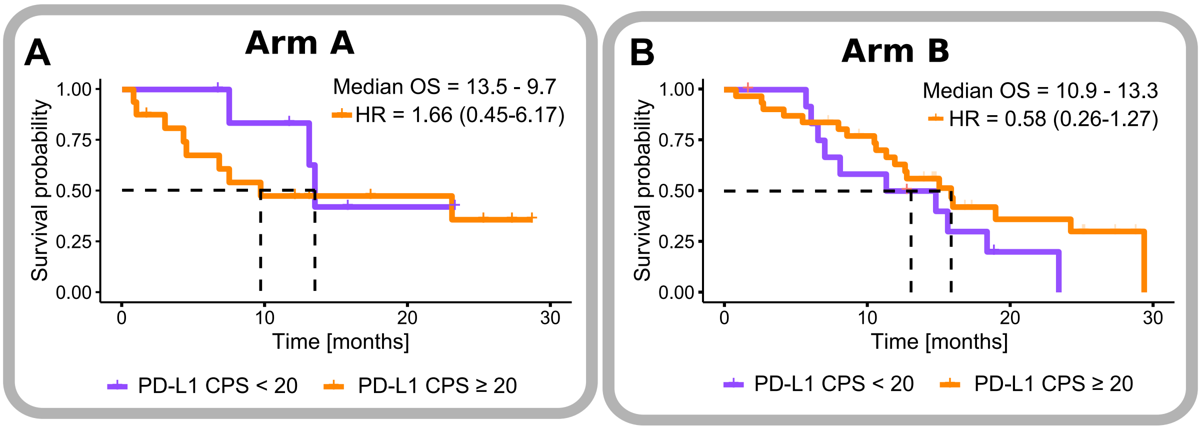


**Supplementary Figure 1: Efficacy of pembrolizumab treatment in patients with R/M HNSCC as a function of PD-L1 CPS (cut-off 20) in the FOCUS trial in the two treatment arms.** **(A)** shows Kaplan-Meier estimates of overall survival (OS) for patients from arm A (without UV-1 vaccination) and **(B)** for patients from arm B (with UV-1 vaccination). HR = hazard ratio. Statistic test = log rank test.

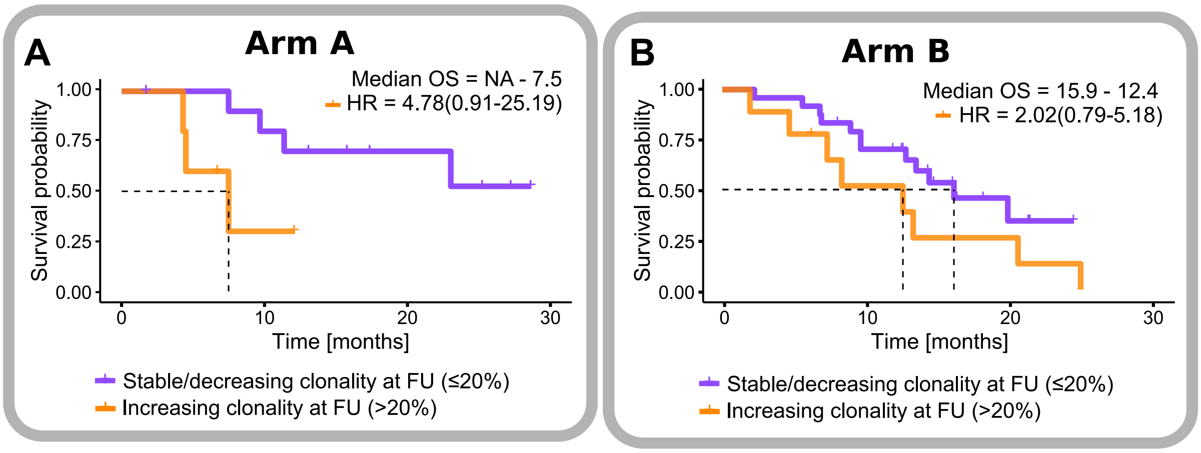


**Supplementary Figure 2: T cell receptor repertoire metrics in HNSCC patients treated with pembrolizumab on the FOCUS trial in the two treatment arms.** Overall survival (OS) outcomes of patients with increasing clonality over the first treatment cycle in arm A (without UV-1 vaccination) **(A)** and B (with UV-1 vaccination) **(B)**. HR = hazard ratio. Statistic test = log rank test.

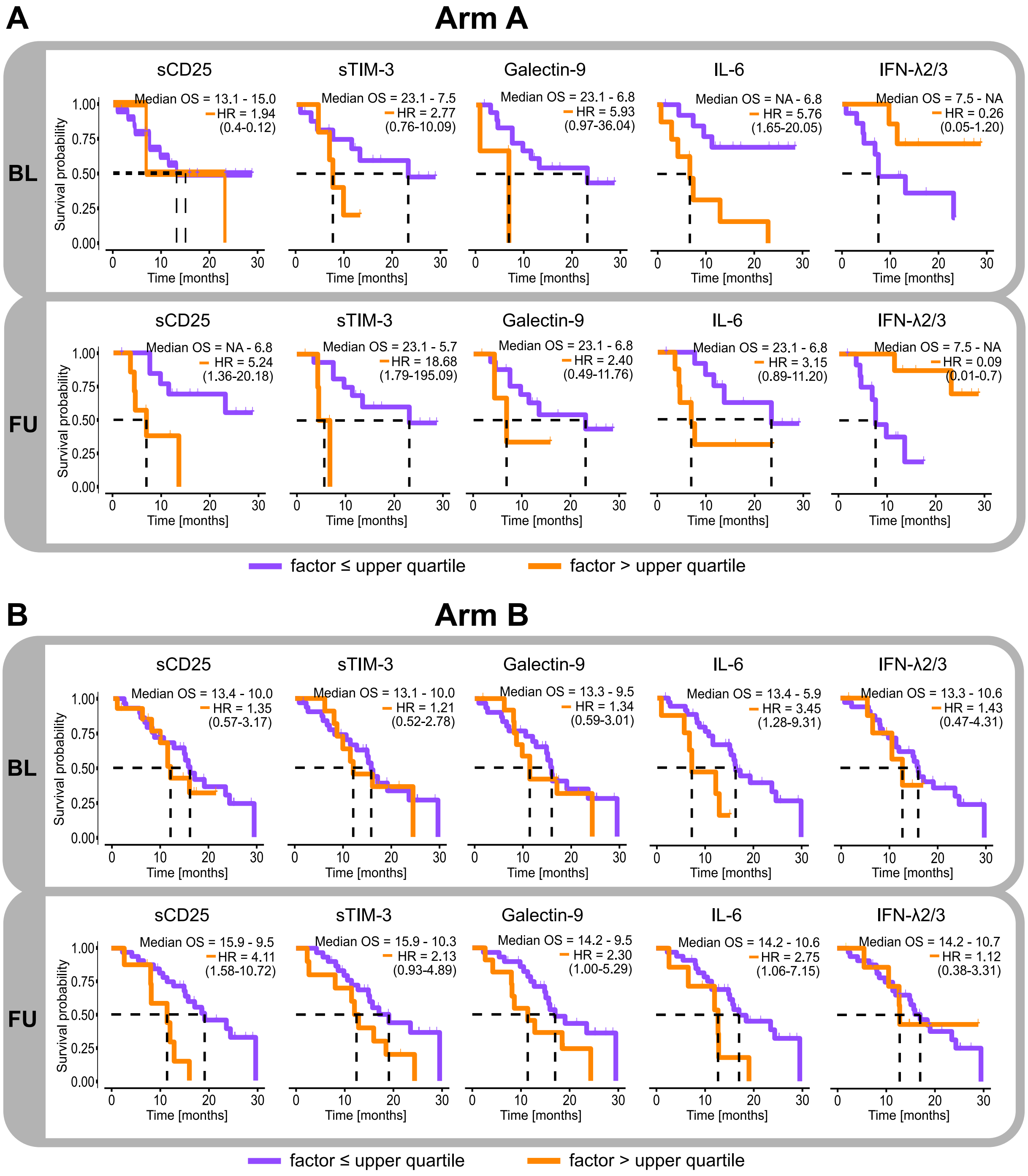


**Supplementary Figure 3: Soluble factor and chemokine profiling baseline (BL) and upon pembrolizumab treatment (FU) in the two treatment arms.** Overall survival outcomes on pembrolizumab in patients with elevated levels of circulating immune checkpoint molecules and cytokines (upper quartile) as compared to the rest of the cohort at BL and at FU in arm A (without UV-1 vaccination) **(A)** and B (with UV-1 vaccination) **(B)**. HR = hazard ratio. Statistic test = log rank test.

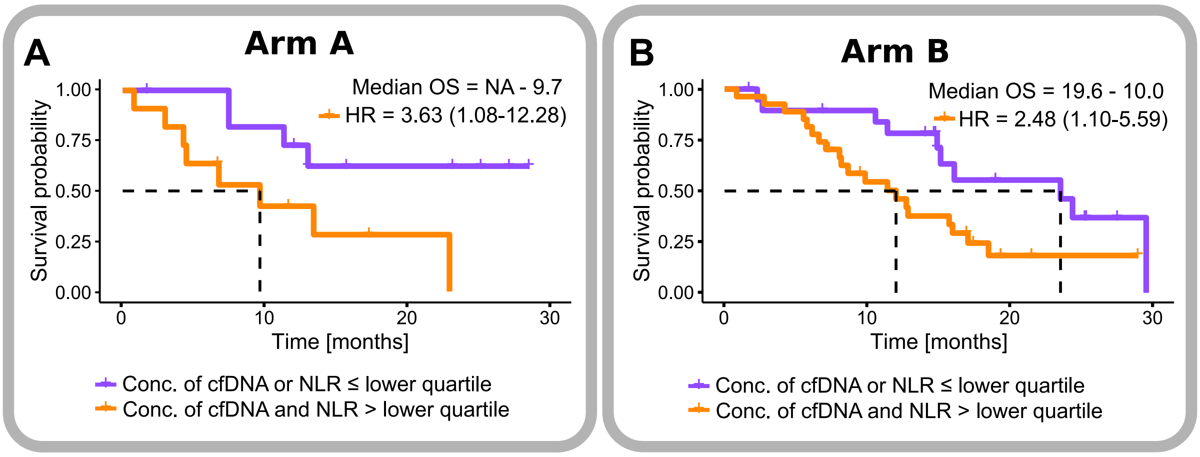


**Supplementary Figure 4: Cell-free DNA (cfDNA) and neutrophil-to-lymphocyte ratios (NLR) at baseline in patients with HNSCC treated with pembrolizumab in the two treatment arms.** Overall survival (OS) in patients with low cfDNA and/or NLR (lower quartile) in arm A (without UV-1 vaccination) **(A)** and B (with UV-1 vaccination) **(B)**. HR = hazard ratio. Statistic test = log rank test.
